# Teaching Uncommon Diseases in Surgery: Conceptual Framework for the Development of a Peritoneal Surface Malignancy Curriculum

**DOI:** 10.1101/2024.04.06.24305431

**Authors:** Frederick A Godley, Varun V Bansal, David G. Su, Vadim Gushchin, Mecker G Moller, Rupen Shah, Alexandra Gangi, Deepa Magge, Dan G Blazer, Oliver S Eng, Laura Lambert, Sean Dineen, Sherif Abdel-Misih, John Encandela, Kiran K Turaga

## Abstract

**Background:** There is a critical need for contemporary education to address peritoneal surface malignancies (PSM). This study delineates the development of an online PSM curriculum for surgical trainees, in conjunction with a national consortium.

**Methods:** A needs assessment survey was administered to attending surgical oncologists and trainees within the consortium, with a focus on current educational practices and preferences for PSM training. The identified focus areas informed the formulation of specific learning objectives and content.

**Results:** The survey was completed by of 86/171 (48.5%) attending surgical oncologists in the group and 70 surgical trainees (56 residents and 14 fellows) from 31 unique institutions. Attending surgical oncologists emphasized trainee familiarity with general PSM principles and peritoneal metastases from lower gastrointestinal and gastric cancers when compared to gynecologic cancers and uncommon primaries (p < 0.001). Attending expectations increased incrementally with the trainee level in the knowledge and patient care domains. Attendings and trainees identified didactics and textbooks as primary modes of learning, although trainees reported using mobile learning tools more frequently. Disease site-specific educational content aligned with learning objectives was uploaded to a previously piloted online learning management system. Clinical management pathways and rotation guides were integrated to enhance the clinical applicability and consistency.

**Conclusions:** Designing a PSM curriculum tailored to the educational needs of both attendants and trainees is feasible by using established pedagogical methods. This study provides a framework for teaching about complex diseases with limited educational literature.

## Introduction

Surgical management of complex medical pathology is continually evolving, and consequently, so is the methodology for teaching and learning about diseases. Effective methods for teaching uncommon, challenging subjects must be engaging and efficiently promote understanding and retention. In recent studies, many attending physicians, trainees, and medical students report insufficient understanding of rare diseases, as well as feeling underprepared to care for patients with rare diseases.^1,2^ Peritoneal surface malignancies (PSMs) represent such clinically significant diseases with rising incidence.^3,4^ Given the concentration of PSM care within academic medical centers^5,6^ the dissemination of expertise remains constrained, hindering access to specialized knowledge.

The multifaceted management of PSM spans perioperative care, surgical principles, and systemic therapy, but resources that comprehensively teach the principles of PSM to surgical trainees are scarce.^7^ Notably, graduates of surgical oncology fellowships in the United States express diminished readiness to manage PSM compared to other oncologic conditions.^8^ Central issues include the absence of standardized recommendations due to inter-institutional PSM practice variability, varying expectations of attending surgeons concerning disease principles across trainee levels, and time constraints limiting trainees’ engagement with traditional learning methods. Consequently, exploring the expectations, perceptions, and educational needs surrounding PSM among attending surgeons and trainees is imperative for optimizing teaching methodologies.

Integrating impressions from these two parallel groups, this study delineates a conceptual framework for developing a standalone online PSM curriculum. The manuscript outlines the curriculum development process in alignment with pedagogical standards, including a targeted needs assessment, followed by devising learning objectives and educational strategies. The ultimate objective is to furnish a model for teaching uncommon surgical diseases, elucidating strategies, and identifying pitfalls encountered during each phase of curriculum development.

## Materials and Methods

This initiative, known as the PSM consortium, is part of a multidisciplinary North American collaborative group process aimed at streamlining care for patients with <u>PSM</u> <u>consortium</u>. The consortium entails a three-part focus: updating management recommendations for PSM based on the foundational Chicago Consensus Guidelines, curriculum development, and delineating institutional standards for PSM care.^9^ The education working group within this consortium, comprising attending surgical oncologists with educational leadership roles and a representative from the Yale Center for Medical Education (co-author JEC), was tasked with developing the PSM curriculum for surgical trainees nationwide. As described herein, Kern’s model for curriculum development was employed, including a targeted needs assessment, followed by outlining learning objectives and educational strategies. ^10^

### 1. Needs Assessment

A pivotal phase in Kern’s model, the needs assessment serves as the cornerstone of curriculum development, offering insights gleaned from key stakeholders, namely attending surgical oncologists and surgical trainees (residents and fellows). Our goals encompassed:

*1. Clarifying Attending Expectations:* Evaluating attending surgical oncologists’ expectations from trainees regarding familiarity with PSM principles across distinct disease sites. These encompassed general PSM, lower gastrointestinal (GI) including colorectal and appendiceal cancers, gastric, gynecological, and uncommon histologies, each subdivided into basic disease principles, surgical care, and systemic therapy.
*2. Defining Clinical Competency Levels:* Identifying the levels of training necessary, as perceived by attending surgeons, to attain competency in various facets of clinical care for PSM patients during the surgical oncology service. These included clinical workflow and electronic health record (EHR) orders, operative principles, perioperative care, national guidelines, research awareness, and palliative care considerations.
*3. Assessing Teaching Modalities:* Comparing the teaching and learning modalities employed by attending surgeons and trainees.
*4. Determining Curriculum Uptake Factors:* Highlighting factors deemed valuable by trainees for curriculum uptake.

#### Survey design and distribution

A cross-sectional survey aligned with the outlined objectives was crafted by the education working group and distributed during a national consortium group meeting in March 2023 via Qualtrics^XM^ (**Appendix**). The survey comprised Likert scale-based items, multiple-choice questions, and free text options, adhering to best practices delineated by Nikiforova et al.^11^ Internal validation was conducted by three attending surgical oncologists in the working group, five surgical residents, and one surgical oncology fellow. While the attending survey was circulated among the consortium group invitees, trainees were accessed through social network/snowball sampling. No response rate was ascertainable because of this strategy. Trainees were encouraged to share the survey with colleagues at their institutions. The survey was deemed exempt by the Yale New Haven Hospital Institutional Review Board.

#### Scoring and Data Analysis

Categorical variables were reported as percentage or frequency responses, whereas continuous variables were reported as median values. Familiarity for trainees has been used as an alternative to competence/proficiency when standardization of trainee assessment is not possible, as in this case.^12^ Trainee familiarity scores were generated by summating the Likert scale responses across related domains, with “not familiar at all” scoring 0, “somewhat unfamiliar” scoring 1, “somewhat familiar” scoring 2, and “very familiar” scoring 3. An identical methodology was used for questions in the trainee survey, gauging self-reported baseline familiarity. As described above, five disease sites were queried, each subdivided by three core principles, with each subdivided topic having a score out of three. Therefore, each disease site had a maximum score of nine, with the maximum overall score across PSM topics being 45. Pairwise comparisons and correlation tests were used to assess the expected familiarity across disease sites and the overall familiarity across training levels.

### 2. Learning objectives

The focus areas identified through the attending needs assessment guided the creation of specific learning objectives for each disease site. These objectives underwent an iterative review by members of the working group and served as the foundational framework for curating educational content.

### 3. Educational content and strategies

The educational content was stratified into three categories. First, written content aligned with the identified objectives was developed utilizing natural language processing (ChatGPT 4) to streamline content creation (**Prompt Template in Appendix**). This content was reviewed and edited by working group members to ensure alignment with the supporting evidence. Second, the updated clinical management pathways devised by the consortium group were integrated with the respective disease sites, enhancing the practical applicability of the educational material. Third, institutional protocols governing PSM management were incorporated. The synthesized content was organized into modules and uploaded onto an online learning management system accessible across universities (Canvas), ensuring compatibility with both computers and mobile platforms.

## Results

### Needs Assessment

A total of 86 of 171 (48.5%) attending surgical oncologists in the consortium and 70 surgical trainees (residents and fellows) from 31 distinct institutions completed the survey. The baseline characteristics of the participants are presented in Tables 1a and 1b. Predominantly, attending surgical oncologists practiced in hospital/medical group settings (96.5%) and university-based centers (77.9%), conducting a median of 25 cytoreductive surgery (CRS) and hyperthermic intraperitoneal chemotherapy (HIPEC) cases annually (median, 25; interquartile range (iQR), 15-40).

**Table 1a:** Attending Demographic Information (n=86). CRS: Cytoreductive Surgery; HIPEC: Hyperthermic Intraperitoneal Chemotherapy; PSM: Peritoneal Surface Malignancy.

|  | **n (% of all respondents)** |
| --- | --- |
| **Structure of Practice** |  |
| Hospital/Medical Group | 83 (96.5%) |
| Private | 2 (2.4%) |
| Government/Military | 1 (1.2%) |
| **Setting of Practice** |  |
| University Center | 67 (77.9%) |
| University Affiliate | 10 (11.6%) |
| Community Center | 9 (10.5%) |
| **Presence of Surgical Oncology Fellowship** | 32 (37.2%) |
| **Presence of Dedicated PSM Service Line** | 55 (64.0%) |
| **Median Number of overall cases performed over the last year** | 150 (95-200) |
| **Median Number of CRS-HIPEC cases performed over the last year** | 25 (iQR 15-40) |

**Table 1b:** Trainee Demographic Information (n=70). Percentages for interest after general surgery include resident level respondents only (n=56).

| **PGY Level** | **n (%)** |
| --- | --- |
| 1 | 3 (4.3%) |
| 2 | 16 (22.9%) |
| 3 | 20 (28.6%) |
| 4 | 5 (7.1%) |
| 5 | 12 (17.1%) |
| Fellow | 14 (20%) |
| **Interest After Surgical Residency** |  |
| Complex General Surgery Oncology Fellowship | 28 (50.9%) |
| Other Fellowship | 18 (32.7%) |
| General Surgery Practice | 5 (9.03%) |
| Other | 4 (7.25%) |

#### Expected familiarity and competency

Attending expectations of trainee familiarity with general PSM principles and lower GI disease sites were significantly higher than those of gastric cancer, which in turn was prioritized over gynecological and uncommon histologies (p < 0.001) (**Figure 1a-b**). Within these domains, greater emphasis was placed on basic disease and surgical principles than on systemic therapies (**Figure 1a**). Overall, expected familiarity across disease sites exhibited a positive correlation with increasing trainee levels, as demonstrated in **Figure 1a & 1c** (r_s_ = 0.66, p < 0.001). A similar trend was identified in the trainee needs assessment, with PGY4-5 residents reporting higher overall baseline familiarity than PGY1-3 residents (median cumulative score: 25 (iQR 18-30) vs. 18 (iQR 11-34), p = 0.01).

**Figure 1a:**
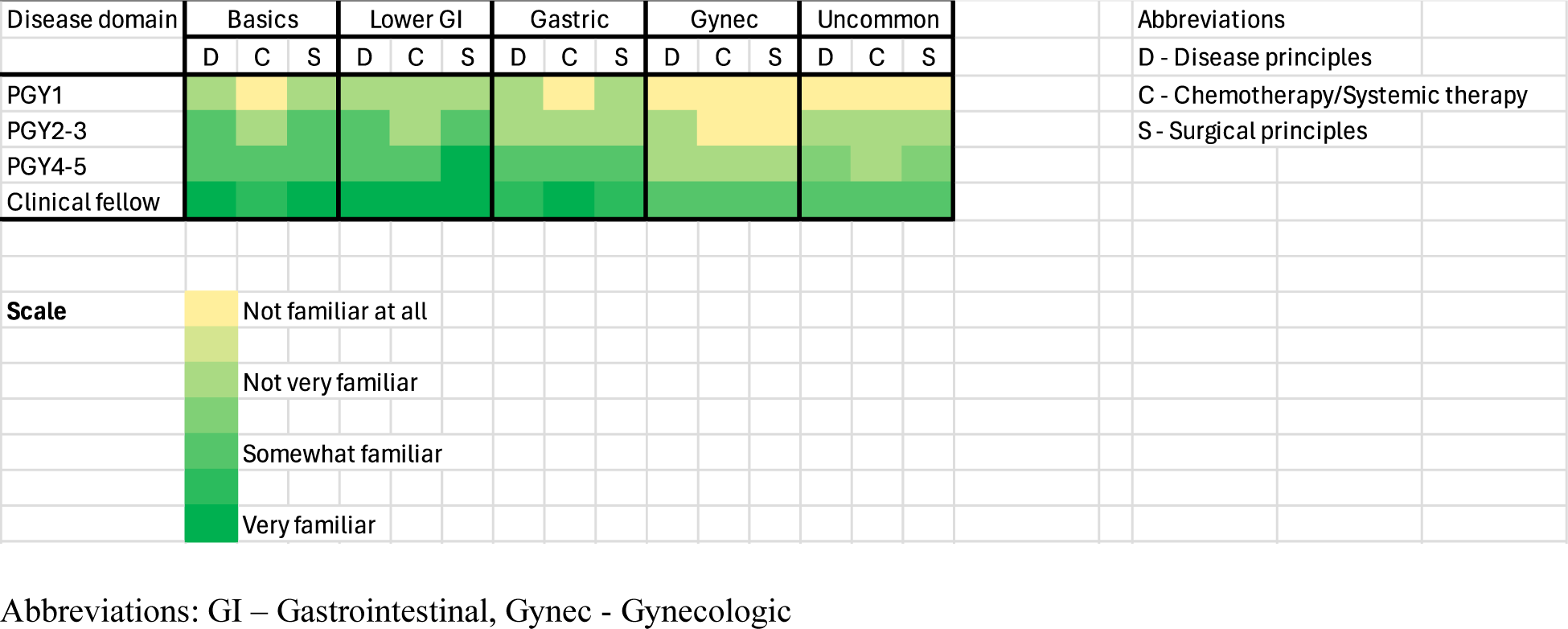
Heat map describing attending expectations for trainee familiarity across disease sites stratified by trainee level.

**Figure 1b:**
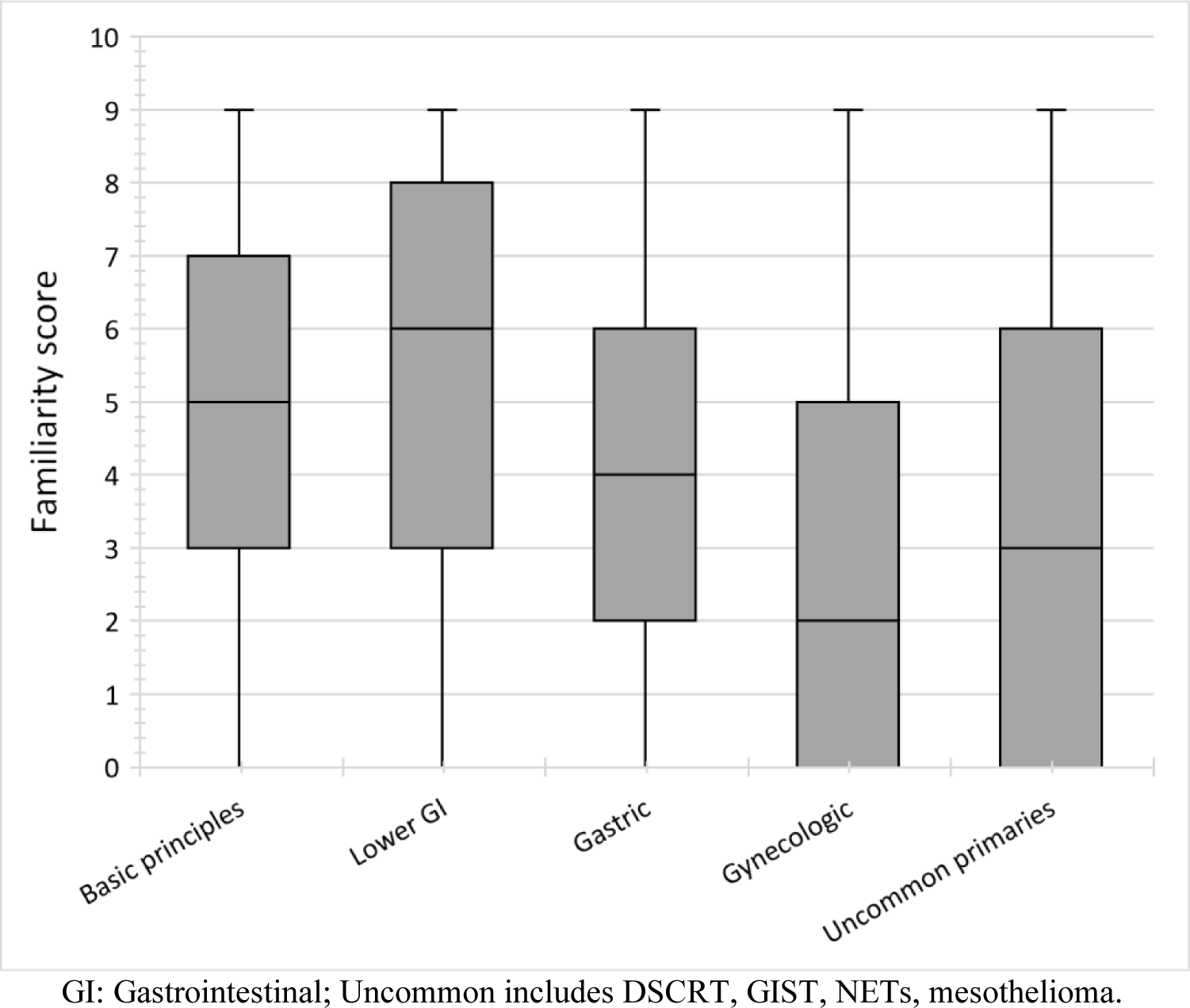
Box and whisker plot demonstrating attending expectations of resident familiarity by disease site. Black horizontal lines represent median familiarity score, while grey bars represent 25-75 percentile of aggregated site scores for general principles, operative management, and systemic therapy (maximum score of 9).

**Figure 1c:**
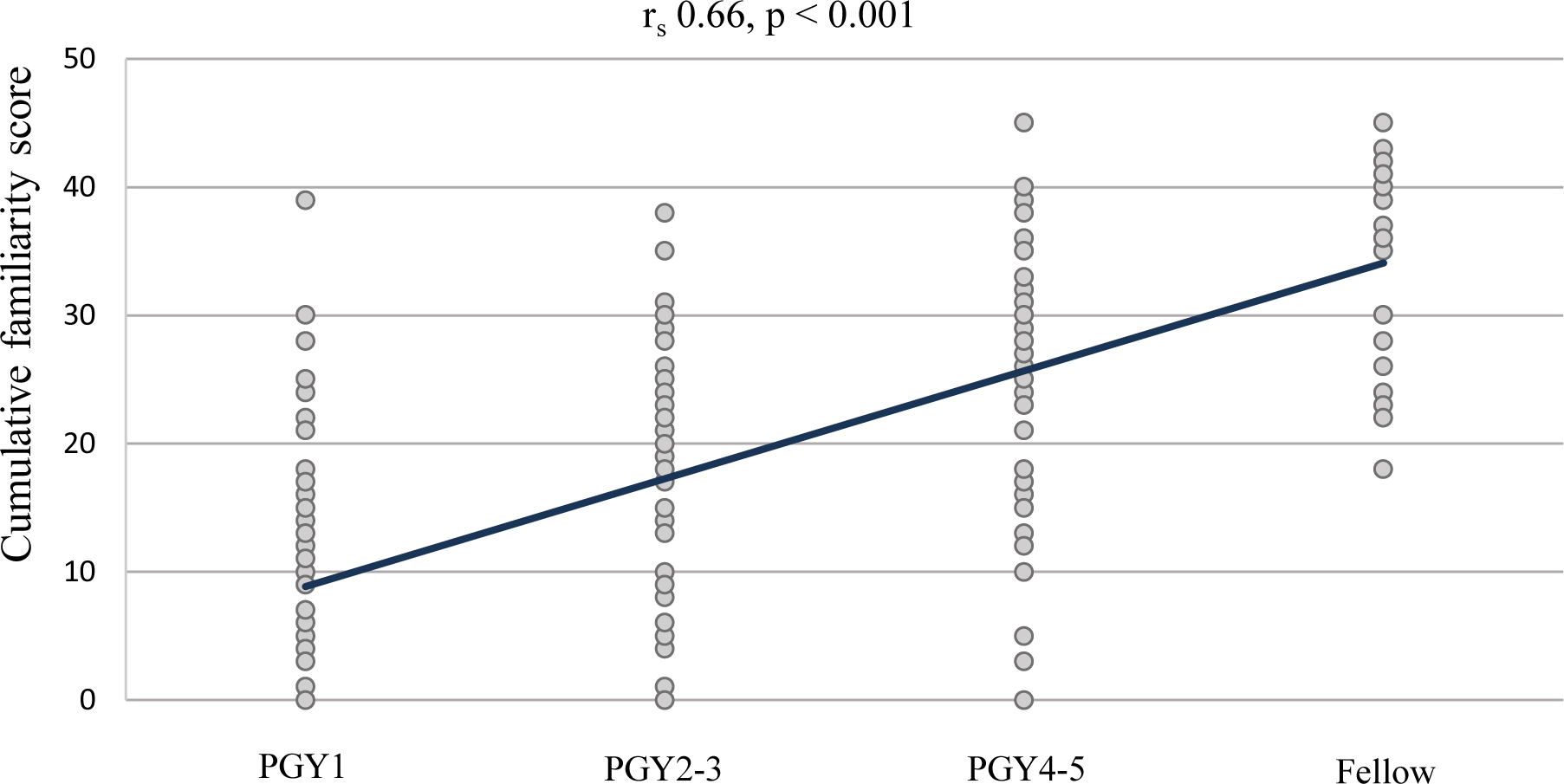
Scatter plot demonstrating attending expectations of cumulative familiarity across disease sites by different levels of training (maximum of 45)

The minimum perceived training levels to achieve competency in PSM care varied: PGY2 (iQR 1-3) for clinical workflow and EHR orders, PGY3 (iQR 2-4) for perioperative management, PGY4 (iQR 3-5) for palliative care considerations, and PGY5 (IQR 4-6/fellow level) for operative principles, treatment algorithms, and research.

##### Educational modalities and curriculum uptake

Both attendings and trainees predominantly relied on didactic (91.9% and 84.3%, respectively) and textbook-based resources (75.6% and 67.1%, respectively) as primary educational modalities. Trainees reported a higher frequency of mobile learning tool usage than attendings (64.3% vs. 50.0%) and also reported using other resources such as question banks and published subject reviews. Concerning protected time for reading while on active surgical service, 72.9% reported having an average of four hours or less per week for studying, only a fraction of which may be available for PSM learning when on the surgical oncology service (**Figure 2**). Concurrently, trainees highlighted concise summaries of important topics (100%), protected time for reviewing content (94.3%), clinical applicability (98.6%), and faculty utilization (85.7%) as crucial factors for the uptake of a dedicated PSM curriculum (**Table 2**).

**Figure 2:**
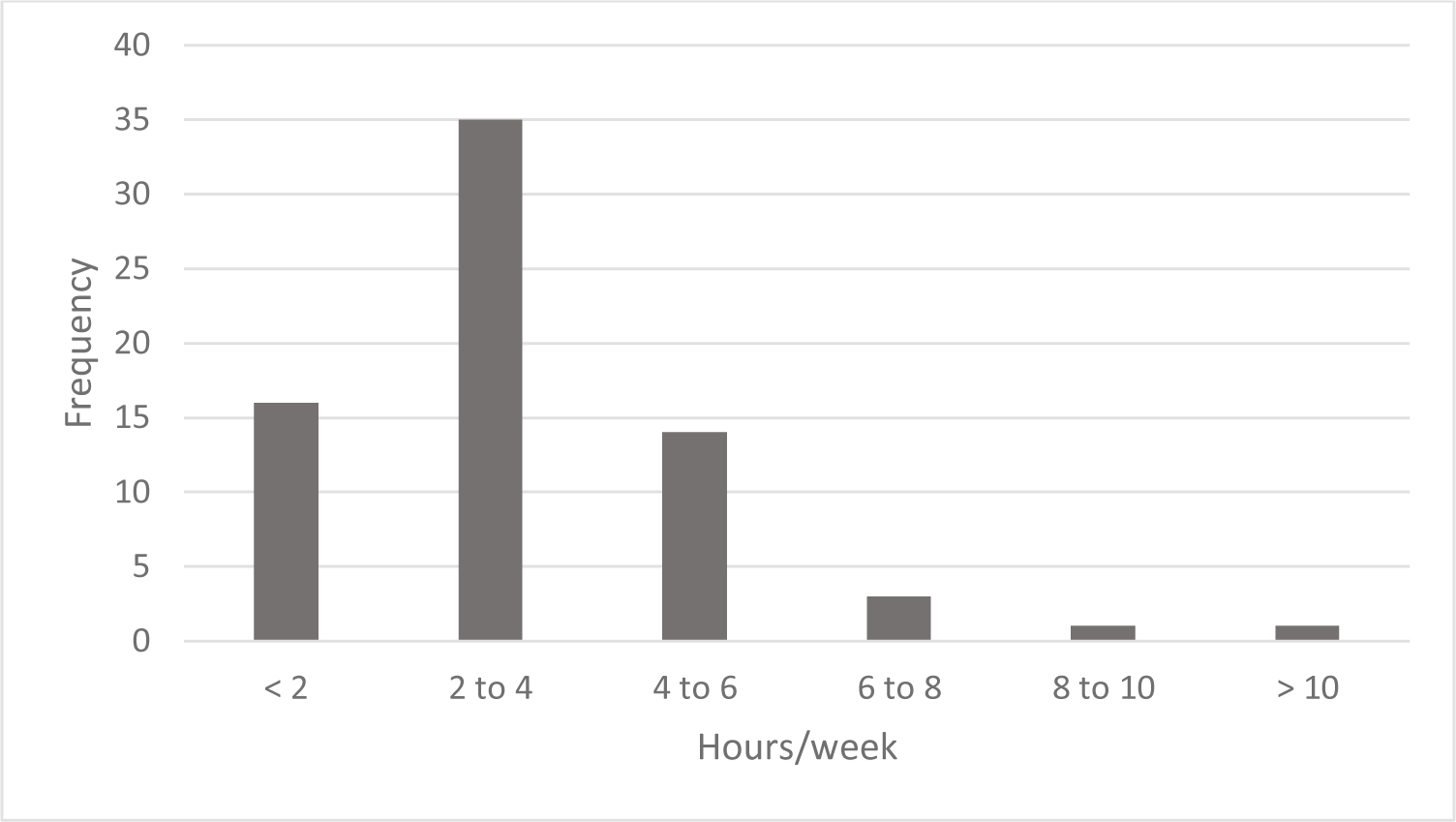
Trainees self-reported time spent on independent study when on clinical rotations.

**Table 2:** Trainee reported importance of factors which they consider important to a PSM course which may be made available to them.

|  | **Frequency of factor cited as “somewhat important” or “very important”: n (%)** |
| --- | --- |
| **Protected Time** | 66 (94.3) |
| **Concise Summary** | 70 (100) |
| **Utilization by Faculty to teach** | 60 (85.7) |
| **Not affect formal evaluations** | 52 (74.3) |
| **Credit for taking the course** | 17 (24.2) |
| **Clinical Applicability** | 69 (98.6) |

### Learning objectives and educational strategies

The attending needs assessment helped delineate focus areas for the curriculum content. Specific learning objectives were written in line with these focus areas, as summarized in **Table 3** and detailed in the **Appendix**. These were further segregated based on the expectations for familiarity and competency elucidated above into core objectives for all learners, and advanced objectives tailored for more experienced learners. The demarcation of training levels for this purpose was informed by the findings of the trainee needs assessment (PGY1-3 vs. 4-5 and fellow). Written content was generated and critically reviewed in accordance with this guiding rubric, and subsequently organized into disease-site-specific modules. Each module was structured to include an 8-to 10-point summary, followed by sections delineating specific learning objectives with anticipated reading time per section, accompanied by links to supplemental audiovisual materials for further comprehension.

**Table 3:**
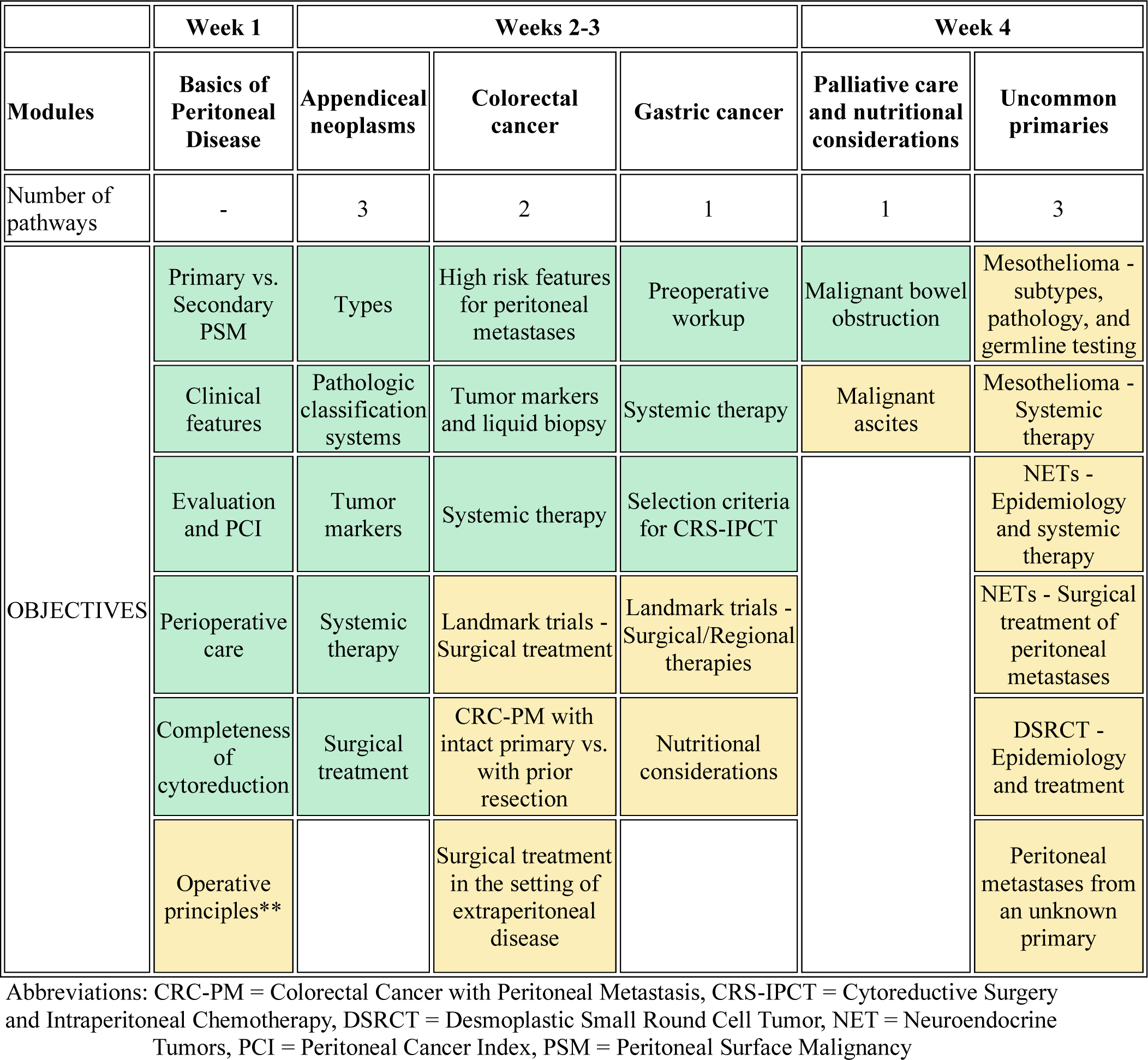
Outline of PSM curriculum modules with subdivisions for course objectives. Core learning objectives (created for trainees PGY1-3) are indicated in green, with advanced learning objectives (for PGY 4-5, clinical fellows) are indicated in yellow. Course objectives were structured using results from needs assessment steps.

To augment the clinical applicability of written content, two significant enhancements were introduced. First, updated consensus-based management pathways for relevant disease sites developed through the collaborative efforts of the PSM consortium were incorporated. These included three pathways for appendiceal tumors (mucinous neoplasms, adenocarcinoma, and peritoneal metastases), two for colorectal cancer (synchronous and metachronous peritoneal metastases), and one each for gastric cancer, peritoneal mesothelioma, neuroendocrine neoplasms, desmoplastic small round cell tumors, and malignant gastrointestinal obstructions. Second, a dedicated module was introduced to provide institution-specific rotation guides, fostering utilization by attending surgeons. These resources comprised EHR ordersets, recommendations for cytoreductive surgery and hyperthermic intraperitoneal chemotherapy (CRS-HIPEC) apparatus setup, enhanced recovery after surgery (ERAS) pathways, and perioperative care protocols tailored to individual attending preferences.

## Discussion

Given the scarcity of literature on PSM within major surgical educational texts, limited mostly to chapter excerpts, this initiative represents the first PSM curriculum tailored for surgical trainees in the United States. The overarching objective was to consolidate nuanced content about PSM into a single, easily accessible online platform, aligning with attendings’ expectations and care preferences and facilitating review by surgical trainees amidst their limited dedicated learning time. The methodology adhered to well-described standards for needs assessments and objective setting across stakeholders.^13–15^

A notable strength of this study lies in its ability to survey both trainees and attendings cross-sectionally through a collaborative group, a best practice for identifying areas of improvement in surgical education. Understanding the needs of both stakeholders is crucial to prevent discordance in the busy clinical setting, fostering an effective learning environment.^16^ In our needs assessment, attendings prioritized general principles and gastrointestinal (GI) cancers, likely reflecting the patient populations encountered in their clinical practice.^17–19^ Notably, both attendings and trainees reported incremental expectations and baseline familiarity with training levels. Attendings consistently expected trainees to be proficient in the clinical and inpatient care of PSM patients as junior residents, while chief residents and surgical oncology fellows were expected to integrate operative principles, palliative care, and national guidelines into their practice when caring for PSM patients.

These expectations collectively helped outline course objectives, facilitating trainees’ transition to more complex levels of learning as they advance through training. Objective delineation offers clarity for learners at various training stages, akin to the Surgical Council for Resident Education (SCORE) curriculum layout.^7^ Moreover, the objectives are aligned with existing international standards for PSM learning as stratified by relevant disease sites in the European Society of Surgical Oncology (ESSO) curriculum.^20^ These objectives were translated closely to clinically applicable domains across institutions by incorporating guidelines and rotation guides collated through the PSM consortium.

While both attendings and trainees predominantly relied on textbooks and didactics as the primary learning media, trainees were more inclined toward mobile-based tools. Online learning platforms present a promising avenue to optimize content for mobile devices and enhance accessibility for clinically active surgical residents and fellows.^21,22^ Our group’s previous work demonstrated the feasibility of implementing an online PSM curriculum through Canvas with acceptable levels of learner engagement, albeit in a single institution.^23^ Future plans entail finalizing curriculum content, integrating clinical scenario-based assessments, and creating summary videos of clinical pathways (chalk talks), followed by pitching to relevant national organizations for hosting content on their online educational platforms to ensure sustainability.

This study bears various limitations in its methodology. The response rate for the attending needs assessment fell below 50%, although the 86 respondents represented a substantial proportion of established national experts in the surgical management of PSM, as estimated from the original Chicago Consensus. The absence of gynecologic oncologists in the survey is notable, reflecting a lower emphasis compared to peritoneal metastases from gastrointestinal malignancies. The trainee needs assessment sample size captured only a fraction of trainees nationwide, primarily due to distribution among attendees of PSM consortium meetings or through their supervising attendings, introducing inherent selection bias. Snowball sampling further exacerbated this bias, limiting respondent diversity and precluding robust comparative analysis because of the uneven representation of training levels and ambiguous questionnaire structures. Other stakeholders such as advanced practice providers and physician assistants were excluded because of limited consortium involvement.

Although the methodology condenses core information efficiently and integrates related topics, it may lack sufficient depth for advanced learners. Therefore, further evaluation and iterative feedback are required prior to implementation in a national standard, such as core competency or entrustable professional activities. Additionally, we assessed familiarity rather than the current levels of competency expected from trainees. Familiarity can be a confusing term for survey respondents but has been highlighted as a precursor to evaluating competency in prior studies.^12^ Our data in this regard, though heterogeneous, holds potential in advancing familiarity with PSM topics as learners progress through training. Future studies should explore competency levels to contextualize familiarity assessments and refine course objectives accordingly.

Despite these limitations, this study offers a reproducible methodology for addressing educational gaps in rare surgical diseases, pivotal given the sparse data, and expertise outside large medical centers. A unique feature of our methodology was the ability to leverage large language models to expedite content generation within ethical purviews. Integration of education alongside systematic reviews of disease processes ensures timely and impactful dissemination of knowledge.^24^ The planned curriculum development reflects major treatment pathway updates, parallel to consortium guideline initiatives, aiming to create a national curriculum accessible to trainees across the country. The consortium aims to meet once every five years, representing a natural opportunity to update course content regularly. Future endeavors aim to tailor PSM educational content for internal medicine, hematology/oncology, and palliative care trainees, exemplifying the ongoing commitment to enhancing rare disease education across disciplines.

## Conclusion

This study highlights the need for a concise and clinically applicable peritoneal oncology curriculum that balances faculty expectations and constraints faced by surgical trainees. It also addresses learning modalities in an evolving educational landscape, with growing preferences for mobile-based tools among trainees. Our methodology provides a blueprint for addressing educational needs regarding rare surgical diseases, emphasizing adaptability and evidence-based approaches.

## Supporting information

Supplemental Item 1: Trainee Survey

Supplemental Item 2: Attending Survey

Supplemental Item 3: Learning Objectives

Supplementary Item 4: ChatGPT Prompt

## Data availability

Data from this study may be made available upon reasonable request from qualified medical or scientific professionals, provided that the request aligns with the specified purpose and may involve de-identified individual participant data. Access to the requested data is granted after signing a data-access agreement.

## Abbreviations

PSM: Peritoneal Surface Malignancies
PGY: Post Graduate Year
EHR: Electronic Health Record.
EPA: Entrustable Professional Activity

