## Supplemental Item 1: Trainee Survey for "Teaching Uncommon Diseases in Surgery: Conceptual Framework for the Development of a Peritoneal Surface Malignancy Curriculum"

### Supplementary Item 1: Surgical oncology survey - Trainee version

Start of Block: Default Question Block

**Peritoneal Cancer Training Needs Assessment - Trainees**

**Introduction:**

Peritoneal surface malignancies (PSMs) represent a complex spectrum of tumors disseminated throughout the lining of the abdomen. Educational materials on this topic are limited in current surgical texts, and dedicated curricula targeting specialized learners and are difficult to access remotely. Therefore, our team developed and piloted an online PSM course to aggregate resources for residents and fellows, which we now aim to revise.

Thank you for your willingness to complete this survey that assesses needs and preferences for a PSM course for residents and fellows. We will combine what we learn from your responses with the lessons we gained from a previous pilot training to develop the next iteration of our online course. The survey should take about **5 minutes** to complete, which you should complete in one sitting since you will not be able to return to the survey at a later time to complete it. Your responses are confidential in that we will not share any information in written or oral reports that might indicate your unique identity.

Q1 First, we would like to know your familiarity with the following **PSM principles**. We do not expect you to be necessarily familiar with PSMs, so your your honesty and accuracy in assessing your familiarity will be helpful in designing the course.

(Note that "Not familiar at all" is the default. If that is your answer for any of the items, just leave the choice; otherwise, click on your other answers..)

| | Level of familiarity with the disease | | | | Level of familiarity with systemic therapy | | | | Level of familiarity with surgical treatment | | | |
|---|---|---|---|---|---|---|---|---|---|---|---|---|
| | Not familiar at all (1) | Not very familiar (2) | Somewhat familiar (3) | Very familiar (4) | Not familiar at all (1) | Not very familiar (2) | Somewhat familiar (3) | Very familiar (4) | Not familiar at all (1) | Not very familiar (2) | Somewhat familiar (3) | Very familiar (4) |

| |
|---|
| Peritoneal Disease Basics (e.g., etiology, pathogenesis, treatment) (1) |
| Lower GI (Appendix and Colorectal) metastasis (2) |
| Gastric cancer metastases (4) |
| Gynecologic primaries (Ovarian, Fallopian Tube and Endometrial) (6) |
| Uncommon Histologies (Mesothelioma, GIST, NETs, DSRCT) (12) |

Q2 In the next part of the survey, we'd like to learn your potential interest in an online course on PSMs. You would be given access to the online course throughout the time that you rotate on the Surgical Oncology service and for review thereafter. Access will be strongly encouraged but not made mandatory. Surgical faculty at your institution will curate, monitor, and update the course through the learning management system at your institution. Given this information, we are interested in your responses to the following.

---

Q3 Which of the following media do you use regularly for learning during residency/fellowship? Choose **all** that apply. Please elaborate

- ☐ Textbook (1)
- ☐ Didactics/Lecture/Conference (2)
- ☐ Mobile learning (cell, tablet) (3)
- ☐ Other (please specify) (4) ________________________________________________

---

Q4 How much time do you dedicate per week to reading/learning while on an active surgical service?

- < 2 hours (1)
- 2 to 4 hours (2)
- >4 to 6 hours (3)
- >6 to 8 hours (4)
- >8 to 10 hours (5)
- > 10 hours (6)

---

Q5 What level of interest do you think you would have in participating in such a course? Click the answer that best describes your interest level. (Please note that your response does not commit you to nor eliminate you from future course participation).

- **Not interested** in the course (1)
- **Not likely to have an interest,** but I might look at course content at some point. (2)
- **Reasonable interest,** I would definitely want to learn more about the course then decide on whether to participate (3)
- **High interest,** I very likely would participate in the course when offered (4)

---

Q6 How important do you think it would be for a trainee at your training level to know the following **about PSMs** by the end of the course?

| | Not important (1) | Of limited importance (2) | Somewhat important (3) | Very important (4) |
|---|---|---|---|---|
| Operative principles (1) | ○ | ○ | ○ | ○ |
| Clinic care flow and EHR orders (2) | ○ | ○ | ○ | ○ |
| Perioperative care in line with attending preferences (3) | ○ | ○ | ○ | ○ |
| Treatment algorithms in line with national guidelines (4) | ○ | ○ | ○ | ○ |
| Landmark studies and active research protocols (5) | ○ | ○ | ○ | ○ |
| Palliative care needs for patients with PSMs (6) | ○ | ○ | ○ | ○ |

Q7 How important do you think the following mechanisms are in maintaining the interest of a trainee at your training level to engage in a course like ours?

| | Not important (1) | Of limited importance (2) | Somewhat important (3) | Very important (4) |
|---|---|---|---|---|
| Protected time for course review (1) | ○ | ○ | ○ | ○ |
| Concise summaries of core information (i.e., despite whether protected time is offered or not) (2) | ○ | ○ | ○ | ○ |
| Course utilization for teaching by faculty (3) | ○ | ○ | ○ | ○ |
| Assuring that course participation or lack thereof does not affect formal evaluations (4) | ○ | ○ | ○ | ○ |
| Course credit (eg. Certificate, CME credits) (5) | ○ | ○ | ○ | ○ |
| Applicability to clinical practice (6) | ○ | ○ | ○ | ○ |
| Other (please specify) (7) | ○ | ○ | ○ | ○ |

Q8 In this last part of the survey, we'd like to ask a few questions about you. This information is needed for research purposes, and some responses may help us to know how to best organize the course for different types of learners.

(As a reminder, if you have provided responses to this survey for our general inforamtion so we may determine needs for the course but have opted out of having your responses included in the research component of our project, it would still be userful to have your responses to the following. If you've opten out, we will not include these or any of your responses to the survey as part of our research report.)

What is your current clinical PGY level (excluding research years)?

- ○ PGY 1 (1)
- ○ PGY 2 (2)
- ○ PGY 3 (3)
- ○ PGY 4 (4)
- ○ PGY 5 and above (5)
- ○ Clinical Fellow (6)

*Display This Question:*

*If In this last part of the survey, we'd like to ask a few questions about you. This information is... != Clinical Fellow*

Q9 If you are a resident, what is your interest after surgical residency?

- ◯ Fellowship (CGSO) (1)
- ◯ Fellowship (Other, please specify) (2) ________________________________________________
- ◯ General Surgery Practice (3)
- ◯ Not sure (4)
- ◯ Other, please specify (5) ________________________________________________

---

Q10 If you are participating in research during residency, what is your research interest?

________________________________________________________________

---

*

Q11 Please mention the institution at which you are currently pursuing your clinical training

________________________________________________________________

**End of Block: Default Question Block**
