## Supplemental Item 2: Attending Survey for "Teaching Uncommon Diseases in Surgery: Conceptual Framework for the Development of a Peritoneal Surface Malignancy Curriculum"

### Supplementary Item 2: Surgical oncology survey - Attending version

Start of Block: Default Question Block

**Peritoneal Oncology Training Needs Assessment - Attendings**

**Introduction:**

Thank you for your willingness to complete this survey that assesses needs and preferences for a peritoneal oncology course for residents and fellows. We will combine what we learn from your responses with the lessons we gained from a previous pilot training to develop the next iteration of our online course for peritoneal surface malignancies (PSMs). The survey should take about **5-10 minutes** to complete, which you should complete in one sitting since you will not be able to return to the survey at a later time. Your responses are confidential in that we will not share any information in written or oral reports that might indicate your unique identity.

Q1 Do you have a Surgical Oncology Fellowship at your institution?

- ○ Yes (1)
- ○ No (2)

Q2 At your institution, trainees at which of the following levels rotate in the Surgical Oncology/PSM service? (Select as many as applicable)

- ☐ PGY1 (1)
- ☐ PGY2 (2)
- ☐ PGY3 (3)
- ☐ PGY4 (4)
- ☐ PGY5 or above (5)

*Display This Choice:*

*If Do you have a Surgical Oncology Fellowship at your institution? = Yes*

- ☐ Clinical fellow (6)

---

Q3 First, we'd like to know your expectations about the level of familiarity a typical trainee in your oncology rotation should have with the following **PSM principles,** with respect to the level of training.

(Note that "Not familiar at all is the default. If that is your answer for any of the items, just leave the choice; otherwise, click on your other answers..)

**PGY1**

| | Level of familiarity with the disease | | | | Level of familiarity with systemic therapy | | | | Level of familiarity with surgical treatment | | | |
|---|---|---|---|---|---|---|---|---|---|---|---|---|
| | Not familiar at all (1) | Not very familiar (2) | Somewhat familiar (3) | Very familiar (4) | Not familiar at all (1) | Not very familiar (2) | Somewhat familiar (3) | Very familiar (4) | Not familiar at all (1) | Not very familiar (2) | Somewhat familiar (3) | Very familiar (4) |

| | | | | | | | | | | | | |
|---|---|---|---|---|---|---|---|---|---|---|---|---|
| Peritoneal Disease Basics (e.g., etiology, pathogenesis, treatment) (1) | ○ | ○ | ○ | ○ | ○ | ○ | ○ | ○ | ○ | ○ | ○ | ○ |
| Lower GI (Appendix and Colorectal) metastases (2) | ○ | ○ | ○ | ○ | ○ | ○ | ○ | ○ | ○ | ○ | ○ | ○ |
| Gastric cancer metastases (3) | ○ | ○ | ○ | ○ | ○ | ○ | ○ | ○ | ○ | ○ | ○ | ○ |
| Gynecologic primaries (Ovarian, Fallopian Tube and Endometrial) (4) | ○ | ○ | ○ | ○ | ○ | ○ | ○ | ○ | ○ | ○ | ○ | ○ |
| Uncommon Histologies (Mesothelioma, GIST, NETs, DSRCT) (5) | ○ | ○ | ○ | ○ | ○ | ○ | ○ | ○ | ○ | ○ | ○ | ○ |

Q4 **PGY2-3**

| | Level of familiarity with the disease | | | | Level of familiarity with systemic therapy | | | | Level of familiarity with surgical treatment | | | |
|---|---|---|---|---|---|---|---|---|---|---|---|---|
| | Not familiar at all (1) | Not very familiar (2) | Somewhat familiar (3) | Very familiar (4) | Not familiar at all (1) | Not very familiar (2) | Somewhat familiar (3) | Very familiar (4) | Not familiar at all (1) | Not very familiar (2) | Somewhat familiar (3) | Very familiar (4) |

| | | | | | | | | | | | | |
|---|---|---|---|---|---|---|---|---|---|---|---|---|
| Peritoneal Disease Basics (e.g., etiology, pathogenesis, treatment) (1) | ○ | ○ | ○ | ○ | ○ | ○ | ○ | ○ | ○ | ○ | ○ | ○ |
| Lower GI (Appendix and Colorectal) metastases (2) | ○ | ○ | ○ | ○ | ○ | ○ | ○ | ○ | ○ | ○ | ○ | ○ |
| Gastric cancer metastases (3) | ○ | ○ | ○ | ○ | ○ | ○ | ○ | ○ | ○ | ○ | ○ | ○ |
| Gynecologic primaries (Ovarian, Fallopian Tube and Endometrial) (4) | ○ | ○ | ○ | ○ | ○ | ○ | ○ | ○ | ○ | ○ | ○ | ○ |
| Uncommon Histologies (Mesothelioma, GIST, NETs, DSRCT) (5) | ○ | ○ | ○ | ○ | ○ | ○ | ○ | ○ | ○ | ○ | ○ | ○ |

Q5 **PGY4-5**

| | Level of familiarity with the disease | | | | Level of familiarity with systemic therapy | | | | Level of familiarity with surgical treatment | | | |
|---|---|---|---|---|---|---|---|---|---|---|---|---|
| | Not familiar at all (1) | Not very familiar (2) | Somewhat familiar (3) | Very familiar (4) | Not familiar at all (1) | Not very familiar (2) | Somewhat familiar (3) | Very familiar (4) | Not familiar at all (1) | Not very familiar (2) | Somewhat familiar (3) | Very familiar (4) |

| | | | | | | | | | | | | |
|---|---|---|---|---|---|---|---|---|---|---|---|---|
| Peritoneal Disease Basics (e.g., etiology, pathogenesis, treatment) (1) | ○ | ○ | ○ | ○ | ○ | ○ | ○ | ○ | ○ | ○ | ○ | ○ |
| Lower GI (Appendix and Colorectal) metastases (2) | ○ | ○ | ○ | ○ | ○ | ○ | ○ | ○ | ○ | ○ | ○ | ○ |
| Gastric cancer metastases (3) | ○ | ○ | ○ | ○ | ○ | ○ | ○ | ○ | ○ | ○ | ○ | ○ |
| Gynecologic primaries (Ovarian, Fallopian Tube and Endometrial) (4) | ○ | ○ | ○ | ○ | ○ | ○ | ○ | ○ | ○ | ○ | ○ | ○ |
| Uncommon Histologies (Mesothelioma, GIST, NETs, DSRCT) (5) | ○ | ○ | ○ | ○ | ○ | ○ | ○ | ○ | ○ | ○ | ○ | ○ |

Q6 **Clinical fellow**

| | Level of familiarity with the disease | | | | Level of familiarity with systemic therapy | | | | Level of familiarity with surgical treatment | | | |
|---|---|---|---|---|---|---|---|---|---|---|---|---|
| | Not familiar at all (1) | Not very familiar (2) | Somewhat familiar (3) | Very familiar (4) | Not familiar at all (1) | Not very familiar (2) | Somewhat familiar (3) | Very familiar (4) | Not familiar at all (1) | Not very familiar (2) | Somewhat familiar (3) | Very familiar (4) |

| | | | | | | | | | | | | |
|---|---|---|---|---|---|---|---|---|---|---|---|---|
| Peritoneal Disease Basics (e.g., etiology, pathogenesis, treatment) (1) | ○ | ○ | ○ | ○ | ○ | ○ | ○ | ○ | ○ | ○ | ○ | ○ |
| Lower GI (Appendix and Colorectal) metastases (2) | ○ | ○ | ○ | ○ | ○ | ○ | ○ | ○ | ○ | ○ | ○ | ○ |
| Gastric cancer metastases (3) | ○ | ○ | ○ | ○ | ○ | ○ | ○ | ○ | ○ | ○ | ○ | ○ |
| Gynecologic primaries (Ovarian, Fallopian Tube and Endometrial) (4) | ○ | ○ | ○ | ○ | ○ | ○ | ○ | ○ | ○ | ○ | ○ | ○ |
| Uncommon Histologies (Mesothelioma, GIST, NETs, DSRCT) (5) | ○ | ○ | ○ | ○ | ○ | ○ | ○ | ○ | ○ | ○ | ○ | ○ |

Q5 In the next part of the survey, we'd like to learn about the current use of educational media in your oncology rotation as well as your expectations for trainees who may benefit from our course.

---

Q7 Which of the following media do your residents/fellows use? Choose **all** that apply. Please elaborate.

- ☐ Textbook (1)
- ☐ Didactics/Lecture/Conference (2)
- ☐ Mobile learning (cell, tablet) (3)
- ☐ Other (please specify) (4) ________________________________________________

---

Q8 Using the sliders, identify **the minimum trainee level** at which you would expect your resident/fellow to achieve **competency** in the following **PSM principles**?

(In the likely scenario that you would expect clinical fellows to achieve competency in all the below areas by fellowship completion, at what level of training would you have expected a clinical fellow to have first achieved this competency? For example, if you would have expected this competency to be reached within the first year of residency, leave the slider at the default (1) for PGY1. Otherwise, move the slider to the training year in which you would have expected the competency to first be achieved.)

| PGY1 | PGY2 | PGY3 | PGY4 | PGY5 | Clinical Fellow |
|---|---|---|---|---|---|

| | 1 | 2 | 3 | 4 | 5 | 6 |
|---|---|---|---|---|---|---|
| Operative principles () | | | | | | |
| Clinic care flow and EHR orders () | | | | | | |
| Perioperative care in line with attending preferences () | | | | | | |
| Treatment algorithms in line with national guidelines () | | | | | | |
| Landmark studies and active research protocols () | | | | | | |
| Palliative care considerations () | | | | | | |

**Lastly, we would like to ask a few questions about the setting of your practice**

Q9 What is the structure of your practice?

- ◯ Hospital/Medical group (1)
- ◯ Private (2)
- ◯ Government/Military (3)
- ◯ Other (4) ________________________________________________

---

Q10 What is the setting of your practice?

- ◯ University center (1)
- ◯ University-affiliated center (2)
- ◯ Community (3)

---

*

Q11 What was the average number of cases you participated in overall as a lead surgeon over the last year

________________________________________________________________

---

*

Q12 What was the average number of CRS-HIPEC you participated in as a lead surgeon over the last year

________________________________________________________________

Q13 Do you have a dedicated PSM service line within the surgical oncology core at your institution?

- Yes (1)
- No (2)

End of Block: Default Question Block
