## Supplemental Item 3: Learning Objectives for "Teaching Uncommon Diseases in Surgery: Conceptual Framework for the Development of a Peritoneal Surface Malignancy Curriculum"

**Supplementary Item 3: Peritoneal Surface Malignancy Curriculum Learning Objectives**

Version 2.1: Last Updated 4/6/2024

General PSM Principles

1. Describe and compare primary and secondary peritoneal surface malignancy (PSM), while identifying at least six tumor sites which can contribute to PSM.
2. Define the components of a PCI score and identify implications of a high PCI score in the setting of PSM across different primary disease sites.
3. Summarize the critical components of a post-operative care plan for PSM patients, including pain management, fluid administration, and nutrition as part of your plan.
4. Chief/Fellow level - Employ appropriate operative techniques in the treatment of peritoneal surface malignancy. Identify
    a. The salient steps of peritonectomy procedures and the associated anatomy (right hemi-diaphragm, left hemidiaphragm, central tendon, IVC bursectomy, anterior, pelvic, mesenteric, and omentectomy)
    b. The components of a HIPEC perfusion apparatus
    c. Most common intraperitoneal chemo regimens (drug, dosage, duration) used across disease sites, their salient properties in terms of disease control, and major associated adverse events in the intraoperative and postoperative period.
    d. Anatomical considerations and steps for vascular resection/reconstruction in the setting of peritoneal metastases

Lower GI (Appendiceal)

1. Describe at least six different types of primary appendiceal neoplasms, including key differences in their pathologic descriptions.
    a. Higher level – Describe and compare pathologic classification systems for epithelial appendiceal neoplasms and pseudomyxoma peritonei (benign and malignant)
2. Identify at least two tumor markers or liquid biopsy options for pre-operative workup of primary appendiceal carcinoma.
3. Compare the indications for different surgical treatments for primary appendiceal tumors, including appendectomy, right hemicolectomy, and CRS/HIPEC. Include the selection of intraperitoneal chemotherapeutic agent (mitomycin C vs. oxaliplatin based regimen) in your surgical plan.
4. FELLOW: Compare the treatment options and prognosis for mucinous appendiceal neoplasm vs. non-mucinous appendiceal neoplasms

Lower GI (Colorectal)

1. Identify the three most common sites of distant metastases from colon and rectal cancer, respectively.

2. Describe anatomic and pathologic features associated with peritoneal metastases from CRC.
3. Identify at least two tumor markers or liquid biopsy options for pre-operative workup of colorectal cancer with suspicion of peritoneal metastasis.
4. Compare the differences in treatment between PSM from CRC versus appendiceal cancer.
5. Chief/Fellow level – Identify landmark clinical trials investigating the surgical management of CRC-PM; critically appraise the utility of intraperitoneal chemo in the management of
    a. High risk CRC with PM
    b. CRC-PM (low vs. high burden PM)
6. Chief/Fellow level – Compare management strategies for CRC-PM in the presence of an intact primary vs. in cases with prior resection.
7. Fellow level – Describe the selection criteria for CRS in the presence of extraperitoneal metastases and anatomical considerations for resection/ablation of liver mets and mets to distant lymph nodes (para-aortic, pelvic, inguinal)
8. Based on the 2018 Chicago Consensus Guidelines, create a workup plan including imaging and consulting services.

Gastric

1. Define the role of tumor markers, imaging, and staging laparoscopy in gastric cancer.
2. Identify appropriate chemotherapy regimens and their sequence (intraperitoneal and systemic) for patients with
    a. Advanced gastric cancer with negative cytology and no PM
    b. Cytology positive gastric ca without clinically evident PM
    c. GCPM
3. Define selection criteria for CRS-HIPEC for patients with GCPM.
4. FELLOW: Define second line chemotherapy for gastric cancer with peritoneal metastases
5. Create a list of important nutritional considerations for patients with gastric cancer in the post-operative period.
6. FELLOW: Create an operative plan for a patient with advanced gastric cancer 2 cm from the GEJ, with D1 lymph node and peritoneal metastases to the liver.

Uncommon Causes of PSM and palliative care

1. Describe at least three unusual cancer types which cause PSM (MPeM, DSCRT, Gyn) and their epidemiology.
2. Define the three pathologic sub-types of peritoneal mesothelioma and compare their selection criteria for CRS.
3. Fellow level - Identify pathologic markers of peritoneal mesothelioma on IHC and describe the prognostic implications of common somatic mutations and chromosomal aberrations.
4. Fellow level – Germline variants

   a. Describe the frequency of germline variants in pleural and peritoneal mesothelioma.
   b. Compare the clinical parameters and surgical treatment options for patients with and without germline variants.
5. FELLOW: Compare the current neoadjuvant and adjuvant treatments of malignant peritoneal mesothelioma and pleural mesothelioma. What is the current role of immunotherapy such as nivolumab in MPM?
6. Describe the frequency of PM from NETs, risk factors for PM, and indications for curative-intent vs. palliative surgery.
7. Outline a palliative care consultation for a patient with PSM from DSRCT.
8. Malignant bowel obstruction due to peritoneal carcinomatosis
   a. Identify common sites of MBO in the setting of PM from GI and gynecologic primaries and their radiologic signs.
   b. Describe indications and available options for surgical intervention in the absence of an acute abdomen.
9. Malignant ascites
   a. Identify common primary sites associated with malignant ascites (exclude pseudomyxoma)
   b. Compare the considerations for surgical treatment of malignant ascites from gynecologic vs. GI primaries.
10. Carcinoma of unknown origin/primary
    In a patient with incidentally identified peritoneal disease in the absence of aknown primary, identify:
        i. Most common potential primary sources
        ii. pertinent investigations (imaging, tumor markers, endoscopic/laparoscopic).
        iii. Salient treatment approaches in line with primary sources
