## Supplementary Item 4: ChatGPT Prompt for "Teaching Uncommon Diseases in Surgery: Conceptual Framework for the Development of a Peritoneal Surface Malignancy Curriculum"

Supplementary Item 4: ChatGPT Prompt for Outlining a Peritoneal Surface Malignancy Curriculum.

This prompt was crafted for use by the authors to generate an outline for the described PSM curriculum. All content generated using large language models was reviewed and confirmed by the authors, who attest to its accuracy in final form.

1. You are writing content for a curriculum for surgical residents and fellows regarding peritoneal surface malignancies
2. Your task is to generate a summary document in line with the following learning objective [Learning Objective #.#]
3. The 3 steps to complete the task are:
    1. Prepare a coherent and cohesive summary
    2. Add appropriate transition sentences to connect the paragraphs
    3. Use bullet points as appropriate but stick to continuous text mostly
    4. The final result should be concise and informative for surgical residents and fellows
    5. Here are the following constraints
        - It should be under 600 words in length
        - The reading time should be less than 3 minutes
